# The genetic architecture of left ventricular non-compaction reveals both substantial overlap with other cardiomyopathies and a distinct aetiology in a subset of cases

**DOI:** 10.1101/2020.01.03.19015602

**Authors:** Francesco Mazzarotto, Megan H. Hawley, Matteo Beltrami, Leander Beekman, Antonio de Marvao, Kathryn A. McGurk, Ben Statton, Beatrice Boschi, Francesca Girolami, Angharad M. Roberts, Elisabeth M. Lodder, Mona Allouba, Soha Romeih, Yasmine Aguib, A. John Baksi, Antonis Pantazis, Sanjay K. Prasad, Elisabetta Cerbai, Magdi H. Yacoub, Declan P. O’Regan, Stuart A. Cook, James S. Ware, Birgit Funke, Iacopo Olivotto, Connie R. Bezzina, Paul J.R. Barton, Roddy Walsh

## Abstract

**Background:** Left ventricular non-compaction (LVNC) is a condition characterised by trabeculations in the myocardial wall and is the subject of considerable conjecture as to whether it represents a distinct pathology or a secondary phenotype associated with other cardiac diseases, particularly cardiomyopathies. We sought to investigate the genetic architecture of LVNC by identifying genes and variant classes robustly associated with disease and comparing these to other genetically characterised cardiomyopathies.

**Methods:** We performed rare variant association analysis using six different LVNC cohorts comprising 840 cases together with 125,748 gnomAD population controls and compared results to similar analyses with dilated cardiomyopathy (DCM) and hypertrophic cardiomyopathy (HCM) cases.

**Results:** We observed substantial overlap in genes and variant classes enriched in LVNC and DCM/HCM, indicating that in many cases LVNC belongs to a spectrum of more established cardiomyopathies, with non-compaction representing a phenotypic variation in patients with DCM- or HCM-causing variants. In contrast, five variant classes were uniquely enriched in LVNC cases, of which truncating variants in *MYH7, ACTN2* and *PRDM16* may represent a distinct LVNC aetiology. *MYH7* truncating variants are generally considered as non-pathogenic but were detected in 2% of LVNC cases compared to 0.1% of controls, including a cluster of variants around a single splice region. Individuals with *MYH7* truncating variants identified in the UK Biobank and cohorts of healthy volunteers also displayed significantly greater non-compaction compared to matched controls, with 50% meeting the diagnostic criteria for LVNC. Additionally, structural variants (exon deletions) in *RYR2* and missense variants in the transmembrane region of *HCN4* were enriched in LVNC cases, confirming prior reports regarding the association of these variant classes with combined LVNC and arrhythmia phenotypes.

**Conclusions:** We demonstrated that genetic association analysis can clarify the relationship between LVNC and established cardiomyopathies, highlighted substantial overlap with DCM/HCM but also identified variant classes associated with distinct LVNC and with joint LVNC/arrhythmia phenotypes. These results underline the complex genetic landscape of LVNC and inform how genetic testing in LVNC cases should be pursued and interpreted.

## Introduction

Left ventricular non-compaction (LVNC) is an abnormality of the left ventricular myocardial wall characterised by a compacted epicardial layer and a non-compacted endocardial layer with prominent trabeculations and intertrabecular recesses. While hypertrabeculation can be observed in a variety of syndromic genetic diseases and as a result of loading conditions (e.g. pregnancy or intensive athletic exercise where it is presumed not to be pathologic^1,2^), LVNC is defined as a cardiac-restricted condition and is typically diagnosed when the ratio of non-compacted to compacted layer (NC/C) is 2 to 2.3. As LVNC can occur either in conjunction with other cardiac diseases or as an isolated phenotype, its true nature is a matter of much debate and conjecture^3–5^. This is reflected also by the different classifications assigned to LVNC by the American Heart Association^6^ (primary genetic cardiomyopathy) and by the European Society of Cardiology^7^ (unclassified cardiomyopathy).

LVNC is observed in patients with an array of genetic cardiac conditions, including cardiomyopathies, arrhythmias, aortopathies and congenital heart disease^8^, suggesting that it may represent a specific phenotypic trait in the presence of an underlying pathology rather than a distinct genetic cardiomyopathy. Initial reports of genetic variants identified in patients with LVNC supported this theory, as most were in sarcomeric genes associated with hypertrophic cardiomyopathy (HCM) and dilated cardiomyopathy (DCM)^9^. The clinical screening of relatives of LVNC patients often detected features more associated with cardiomyopathies other than LVNC^10^, suggesting that in certain individuals other genetic or environmental factors interact with a cardiomyopathy-predisposing mutation to produce a non-compaction phenotype. However, the true nature and genetic aetiology of LVNC, and whether it can be considered a separate disease entity or a secondary phenotype occurring in certain patients with primary cardiomyopathies and other cardiac diseases, remains uncertain.

Several recently published studies have used large panels of genes associated with inherited cardiac conditions to evaluate the genetic basis of LVNC^11–15^. However, while providing valuable insights, these studies were individually underpowered to establish statistically robust association of individual genes, particularly for genes that are rarely causative. Additionally, the use of large panels that include genes not validated as causative in LVNC increases the risk of false positive associations, particularly where uniform classification criteria are applied for all genes. As with all genetic disease, the identification of fully validated disease-causing genes is a prerequisite for the successful and reliable adoption of genetic testing in the clinic.

The application of candidate gene studies and large panel sequencing to investigate the genetic basis of cardiomyopathies has led to a large expansion in gene to disease associations, though many lacked robust statistical evidence. Large population genetic databases including ExAC and gnomAD (https://gnomad.broadinstitute.org/) have demonstrated an unexpectedly large prevalence of rare variation in many genes, resulting in the need to re-assess these potential associations and highlighting how rarity alone is not sufficient to deem a variant as disease-causing. While stringent assessment of the evidence linking genes to diseases, such as that applied by the ClinGen initiative^16,17^, can highlight erroneous associations, rare variant burden analysis provides a robust statistical approach to identify and prioritise the most relevant genes for Mendelian diseases. We have recently used these techniques to clarify the genetic basis of cardiomyopathies^18,19^, and confirmed that genes characterised by a significant excess of rare variants in cases versus controls account for the vast majority of HCM patients with an identified pathogenic variant^20^.

Here we perform a meta-analysis of four previously published and two unpublished cohorts of sequenced LVNC cases to identify the genes and variant classes significantly associated with this condition. Such genes are likely to account for the preponderance of disease-causing variants in LVNC patients and should therefore be prioritised for genetic testing. By comparing these findings with equivalent data from other cardiomyopathy cohorts, we can determine the extent to which LVNC is a distinct genetic disease or a phenotypic expression of other cardiomyopathies or cardiac diseases. We demonstrate substantial overlap between LVNC and DCM/HCM but also identify variant classes that are distinctly associated with LVNC and LVNC/arrhythmia phenotypes.

## Methods

### LVNC cohorts

Six distinct LVNC cohorts were assessed in this study and are summarised in Table 1 (the number of cases sequenced per gene in each cohort is detailed in Table S1). Two of these cohorts, Careggi University Hospital in Florence, Italy (32 cases) and the Laboratory for Molecular Medicine (LMM), Partners Healthcare, Boston, USA (233 cases) were previously unpublished. All cohorts were from European or North American groups and laboratories and therefore cases were likely to be of majority Caucasian ethnicity. All cohorts were sequenced with broad panels of genes associated with cardiomyopathies and other cardiac conditions, with the exception of the study by Klaassen *et al*.^9,21,22^ (sequencing restricted to 8 sarcomeric genes and *PRDM16*) and Miszalski-Jamka *et al*.^11^ (whole-exome sequencing was performed but only data from 104 genes was published). For previously published studies, we included in our analysis only those studies where all rare variants detected in cases were listed, regardless of their diagnostic classification. For the LMM cohort, data collection for this study was approved by the Partners HealthCare Institutional Review Board (protocol 2006P001108: Genotype-Phenotype Studies for Cardiomyopathies). For the Florence cohort, all participants gave written informed consent and the study was approved by the relevant regional research ethics committee. See Supplemental Methods for full details of each cohort.

**Table 1:**
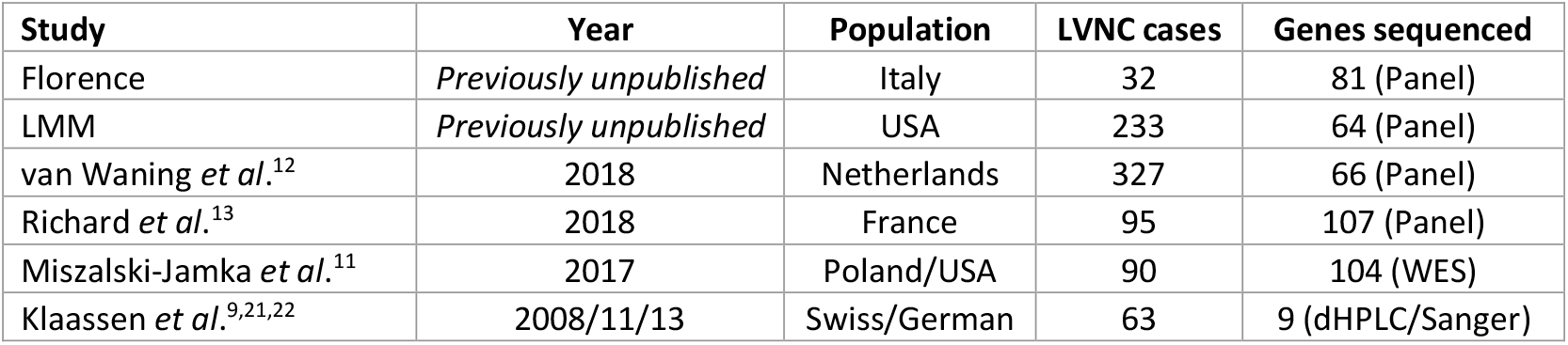
Details of the LVNC cohorts assessed in this study.

| Study | Year | Population | LVNC cases | Genes sequenced |
| --- | --- | --- | --- | --- |
| Florence | *Previously unpublished* | Italy | 32 | 81 (Panel) |
| LMM | *Previously unpublished* | USA | 233 | 64 (Panel) |
| van Waning *et al.*<sup>12</sup> | 2018 | Netherlands | 327 | 66 (Panel) |
| Richard *et al.*<sup>13</sup> | 2018 | France | 95 | 107 (Panel) |
| Miszalski-Jamka *et al.*<sup>11</sup> | 2017 | Poland/USA | 90 | 104 (WES) |
| Klaassen *et al.*<sup>9,21,22</sup> | 2008/11/13 | Swiss/German | 63 | 9 (dHPLC/Sanger) |

### Rare variant burden testing for LVNC cases

Rare variant burden testing between case cohorts and gnomAD exomes (v2.1) population reference samples (n=125,748) was performed as previously described^18^. Rare variants were defined as having a filtering allele frequency (FAF) in gnomAD less than 0.0001 (see Supplemental Methods for further details). Analyses were performed separately for predicted truncating variants (nonsense, frameshift, and splice donor and acceptor variants) and non-truncating variants (missense, small inframe insertions/deletions and stop lost). The frequency of rare variants in 70 genes (i.e. those sequenced and reported in at least half of the constituent disease cohorts to focus on the most relevant genes for LVNC) was compared between LVNC cases and gnomAD. The number of LVNC cases sequenced per gene ranged from 173 to 820. All rare variants detected in LVNC cases were included in burden testing regardless of the clinical classification applied in any of the constituent cohorts. To account for variable coverage in exome-sequenced gnomAD samples, the number of individuals deemed sequenced per gene in gnomAD was calculated using the mean number of alleles across all rare protein-altering variants detected in gnomAD for the gene in question, and then converted from allele number to individuals (divided by 2 for autosomal genes and 1.46 for X chromosome genes based on the male-female ratio in gnomAD). Statistical significance for enrichment of variants in cases was assessed with a one-sided Fisher’s exact test, with Bonferroni multiple testing correction applied for the number of genes tested. The case excess was defined as the difference in rare variant frequencies between case and population reference cohorts. The full list of rare variants detected in each LVNC cohort is listed in Table S2.

For those genes not showing overall enrichment, we tested for potential domain-specific enrichment of non-truncating variants in LVNC cases using an unsupervised, sequence-based clustering algorithm^23^ (details in Table S3). Furthermore, we also assessed variant enrichment in cases for the established *RBM20* DCM pathogenic hotspot between residues 634 and 638. In addition, we analysed the occurrence of structural variants (SVs) in *RYR2* in LVNC cases and controls, based on previously published reports (further details below and in Supplemental Methods).

### Comparison to gene associations in other cardiomyopathies

Variant classes with a significant excess in LVNC versus gnomAD were compared to the results of similar analyses in DCM and HCM cohorts^18,20^ (see Supplemental Methods and Table S4 for details of the cohorts used). Variant classes enriched in LVNC patients as well as in DCM and/or HCM indicate a potential shared genetic aetiology between LVNC and the more well characterised cardiomyopathies, whereas those unique to LVNC suggest a distinct aetiology.

### Effect of MYH7 truncating variants in population controls

The maximum non-compacted to compacted layer (NC/C) ratios in individuals with a *MYH7* truncating variant in UK Biobank (n=12,447 individuals with both exome sequencing and cardiac magnetic resonance imaging) and healthy volunteers from the UK Digital Heart Project (n=912)^24^ and the Egyptian Collaborative Cardiac Genomics (ECCO-GEN) Project (n=400)^25^ were compared to an equivalent number of year-of-birth-, sex-, and ethnicity-matched non-carriers to assess the effect of these variants on non-compaction (Figure S1 and further details in Supplemental Methods). Significance was assessed with a one-sided Wilcoxon Rank Sum test.

## Results

### Rare variant burden in LVNC cases versus gnomAD

To investigate the genes and variant classes associated with LVNC, we compared the frequency of rare variation in six LVNC cohorts to gnomAD exomes population controls. A significant excess of rare variants was seen in LVNC cases compared with gnomAD (p<0.0007 with Bonferroni adjustment for testing 70 genes) in the case of truncating variants in *TTN* (excess burden in cases = 8.6%), *MYBPC3* (2.0%), *MYH7* (2.0%), *PRDM16* (1.3%), *ACTN2* (0.6%) and *RBM20* (0.5%), for non-truncating variants in *MYH7* (10.4%), *ACTC1* (2.0%), *MYBPC3* (1.7%), *TNNT2* (1.6%), *TPM1* (0.8%) and structural variants (SVs, exon deletions) in *RYR2* (1.2%) (Figure 1 and Table 2, for full details on all genes see Table S5). Although non-truncating variants were not significantly enriched overall for *RBM20* and *HCN4*, a significant excess was observed for the DCM pathogenic hotspot in *RBM20* (0.5%) and the transmembrane region of *HCN4* (3.2%). Based on the overall excess in these significant variant classes alone, a causative genetic variant would be identified in an estimated 35.4% of LVNC cases, in line with contemporary estimates for other cardiomyopathies^18,19^.

**Table 2:** Variant classes with a significant excess (after Bonferroni multiple testing correction) of rare variants in combined LVNC cohorts compared to gnomAD exomes. For TTN, only variants affecting exons included in >90% of the transcripts (Percent Spliced In (PSI) >0.9) were included^26^. For HCN4 and RBM20, only variants within the transmembrane region and pathogenic hotspot respectively were included, as described in Methods. For RYR2, only structural variant exon deletions are noted and compared to equivalent variants in gnomAD genomes.

| Gene | Variant Class | Other cardiac associations | gnomAD count/total | gnomAD freq. | LVNC count/total | LVNC freq. | Case excess | p-value | Odds ratio (95% CI) |
| --- | --- | --- | --- | --- | --- | --- | --- | --- | --- |
| MYH7 | Non-Truncating | DCM, HCM | 1754/124979 | 1.40% | 97/820 | 11.83% | 10.43% | 7.7E-56 | 9.4 (7.6-11.7) |
| TTN | Truncating | DCM | 590/122054 | 0.48% | 47/516 | 9.11% | 8.63% | 1.2E-42 | 20.6 (15.1-28.1) |
| ACTC1 | Non-Truncating | DCM, HCM | 73/125274 | 0.06% | 16/760 | 2.11% | 2.05% | 3.1E-19 | 36.9 (21.4-63.7) |
| MYBPC3 | Truncating | HCM | 89/115675 | 0.08% | 17/805 | 2.11% | 2.04% | 1.7E-18 | 28.0 (16.6-47.3) |
| MYH7 | Truncating | - | 97/124979 | 0.08% | 17/820 | 2.07% | 1.99% | 2.4E-18 | 27.3 (16.2-45.8) |
| PRDM16 | Truncating | - | 7/120147 | 0.01% | 6/444 | 1.35% | 1.35% | 4.0E-12 | 235.1 (78.7-702.4) |
| TNNT2 | Non-Truncating | DCM, HCM | 227/124805 | 0.18% | 13/741 | 1.75% | 1.57% | 2.8E-09 | 9.8 (5.6-17.2) |
| HCN4 (tm) | Non-Truncating | Bradycardia | 107/103942 | 0.10% | 7/214 | 3.27% | 3.17% | 4.8E-09 | 32.8 (15.1-71.4) |
| RYR2 | Exon deletion | CPVT | 0/10738 | 0.00% | 5/429 | 1.17% | 1.17% | 8.2E-08 | 278.3 (15.4-5040.7) |
| ACTN2 | Truncating | - | 13/125085 | 0.01% | 4/611 | 0.66% | 0.65% | 1.3E-06 | 63.4 (20.6-195.0) |
| RBM20 (hs) | Non-Truncating | DCM | 1/76260 | 0.00% | 3/546 | 0.55% | 0.55% | 1.4E-06 | 421.3 (43.8-4056.9) |
| TPM1 | Non-Truncating | DCM, HCM | 105/124430 | 0.08% | 6/702 | 0.85% | 0.77% | 4.2E-05 | 10.2 (4.5-23.3) |
| RBM20 | Truncating | DCM | 18/76260 | 0.02% | 3/546 | 0.55% | 0.53% | 4.3E-04 | 23.4 (6.9-79.7) |
| MYBPC3 | Non-Truncating | HCM | 1780/115675 | 1.54% | 26/805 | 3.23% | 1.69% | 4.6E-04 | 2.1 (1.4-3.2) |

**Figure 1:**
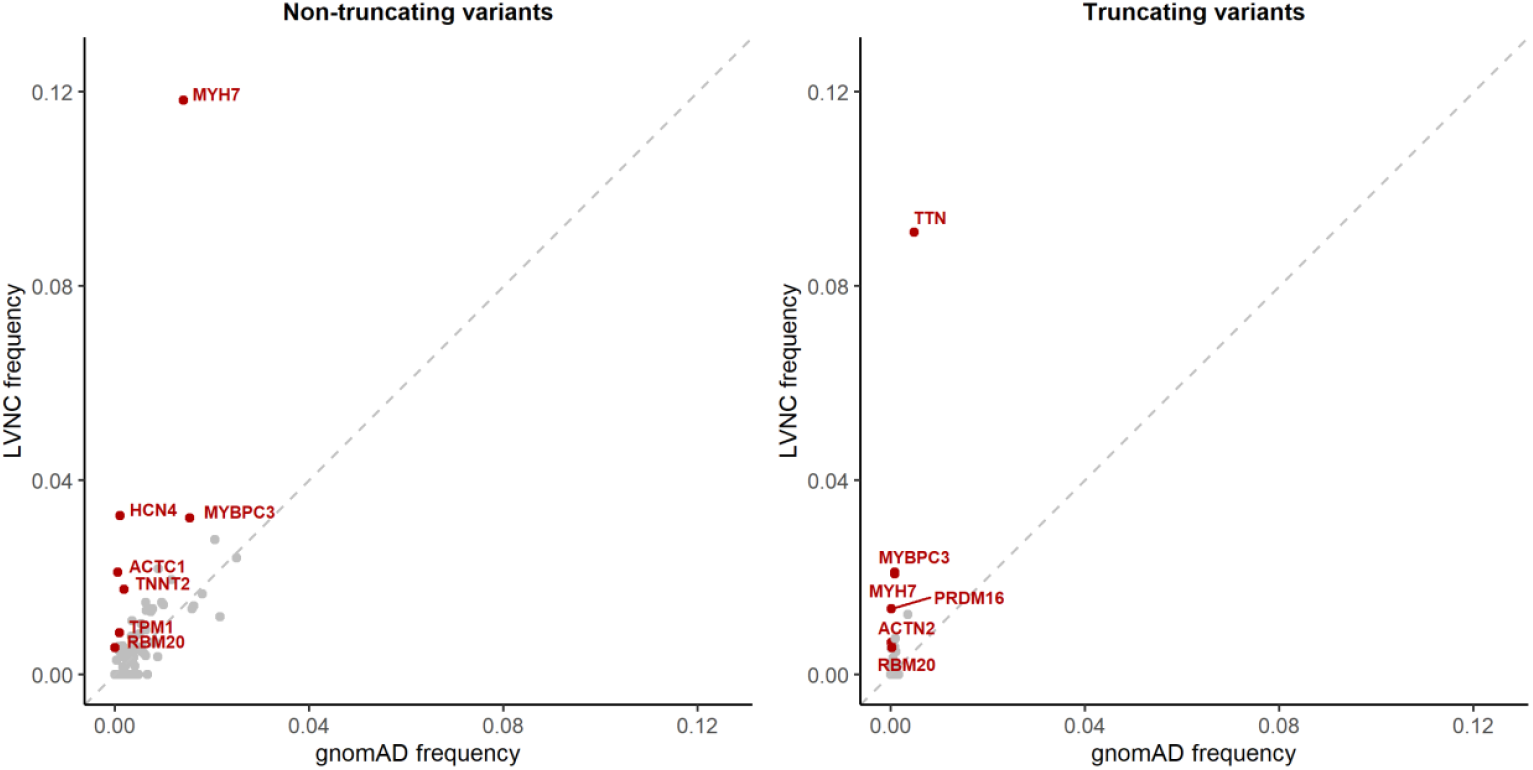
Comparison of the frequency of rare variants for the combined LVNC cohorts (y-axis) and gnomAD population reference samples (x-axis), for non-truncating variants (left) and truncating variants (right). Genes with a significant excess in LVNC cases (p<0.0007 with multiple testing correction) are highlighted in red. For non-truncating variants, data is restricted to the transmembrane region and pathogenic hotspot for HCN4 and RBM20 respectively, as described in the text.

### Overlap with variant classes associated with HCM and DCM

The variant classes enriched in LVNC were compared to those enriched in DCM and HCM (Figure 2), to enable assessment of the genetic overlap with LVNC. Truncating variants in *TTN* and *RBM20*, as well as non-truncating variants within the pathogenic DCM hotspot of *RBM20*, are significantly enriched in both LVNC and DCM. Truncating and non-truncating variants in *MYBPC3* are enriched in both LVNC and HCM. The proportion of LVNC cases with variants in *TTN* and *MYBPC3* is notably lower compared to DCM and HCM respectively, which may reflect the more heterogeneous aetiology of LVNC (Figure 2). A significant excess of non-truncating variants in four other sarcomeric genes (*MYH7, TNNT2, TPM1, ACTC1*) is observed in all three conditions. Of the enriched variant classes, non-truncating variants in *MYH7* had the highest frequency in LVNC cases (11.8%). These variants are also commonly observed in DCM and HCM (Figure 2). However, distinctive (though overlapping) patterns of variant clustering were observed in LVNC and HCM cohorts (Figure 3A), with variants in LVNC cases clustered around the N-terminus myosin head region (residues 39-415).

**Figure 2:**
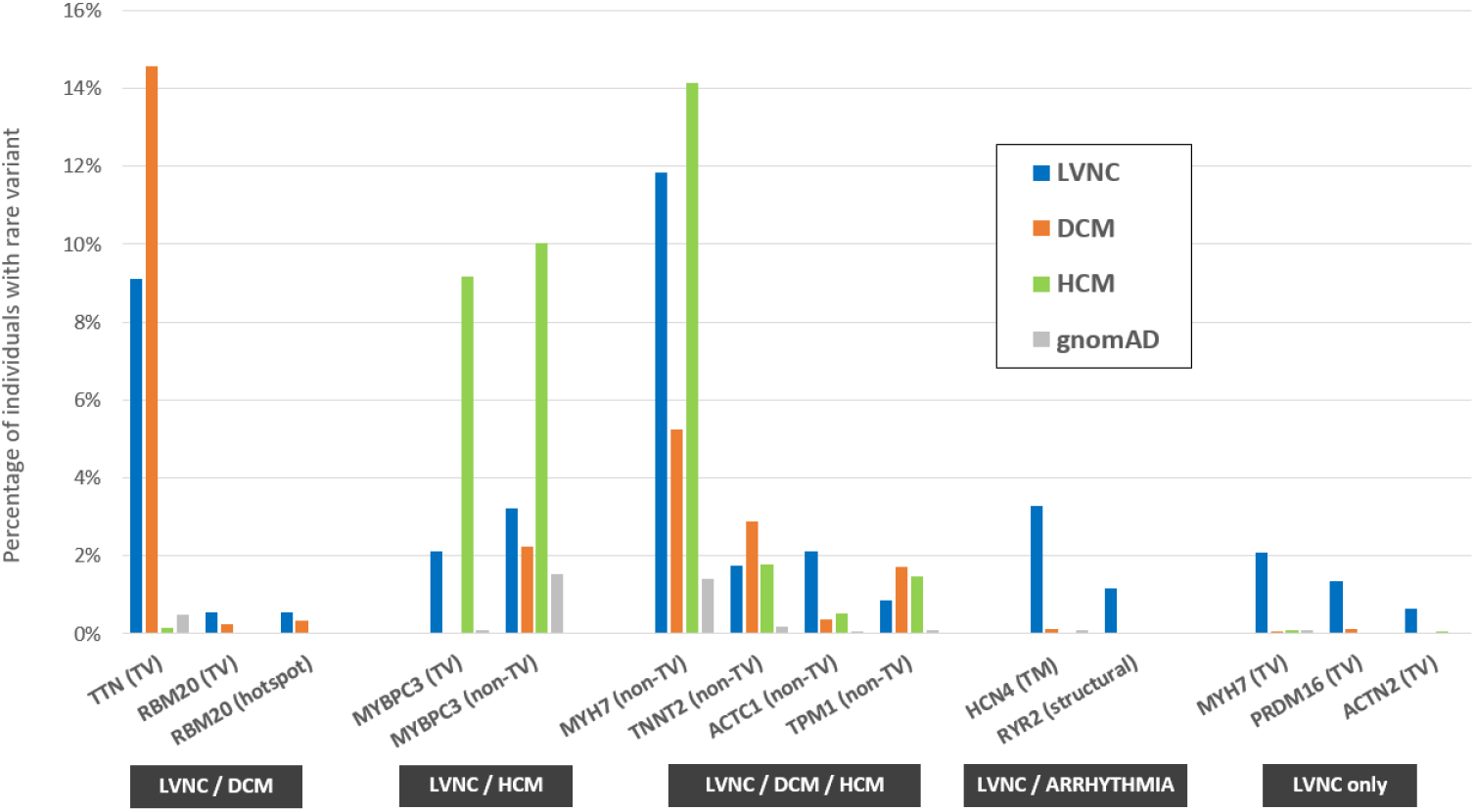
Variant classes with a significant excess in LVNC cases versus gnomAD, with comparison to equivalent frequencies of rare variants in DCM and HCM cohorts. Variant classes are grouped according to which conditions display significant enrichment over gnomAD – LVNC and DCM, LVNC and HCM, all three indications, LVNC and arrhythmia phenotypes and LVNC only. Variant classes shown are truncating variants (TV), non-truncating variants (non-TV) and pathogenic hotspot for RBM20, transmembrane region (TM) for HCN4 and structural variants for RYR2.

**Figure 3:**
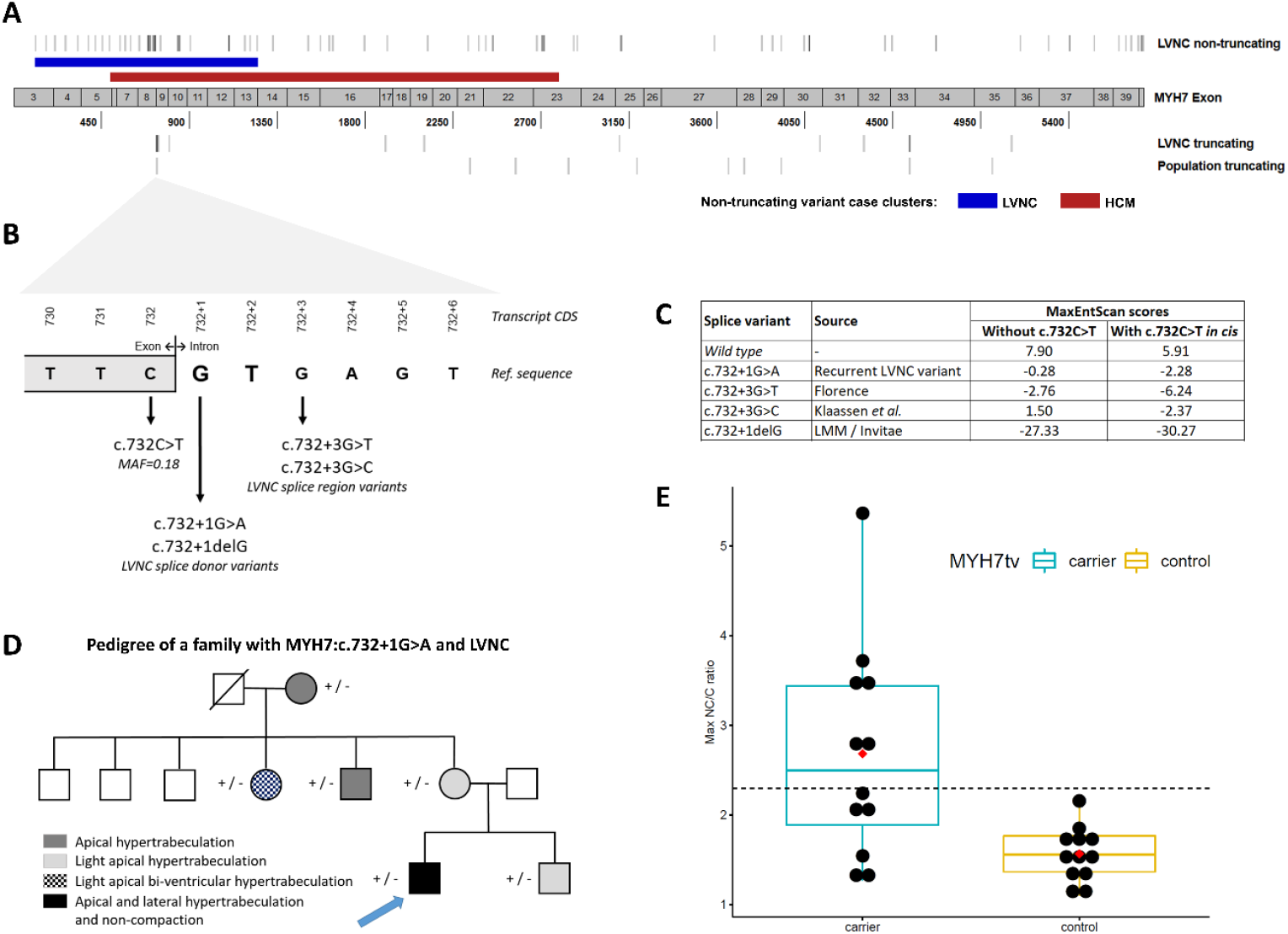
(A) Distribution of rare MYH7 non-truncating variants demonstrates distinct (though overlapping) enriched clusters in LVNC cases (blue band) and HCM cases^23^ (red band). MYH7 truncating variants are distributed throughout the transcript in LVNC cases and population cohorts but with a cluster around the c.732 splice region. (B) Details of the c.732 splice region and associated variants found in LVNC cases. (C) MaxEntScan^28^ scores for these variants (and the wild type sequence) with the reference exon base at c.732 and the c.732C>T common variant in cis. (D) Pedigree of the Italian family demonstrated segregation of the c.732+1G>A variant with non-compaction and/or varying degrees of myocardial hypertrabeculation. The proband alone additionally carried the PRKAG2:c.247C>T variant which, although classified as “likely benign” due to its frequency in population databases (0.13% in South Asians in gnomAD), has been proposed as a modifier aggravating the HCM phenotype in a small pedigree^29^ and therefore may contribute to the more severe phenotype observed in the proband. (E) MYH7-truncating variants are associated with LV non-compaction in population cohorts. Maximum NC/C ratios of individuals identified with MYH7-truncating variants in population controls not selected for disease (see Methods), compared with age-and sex matched individuals without such variants drawn from the same populations. Boxplots show the median and interquartile range, red diamond indicates mean and the dashed line shows the diagnostic NC/C ratio of 2.3.

### Variant classes unique to LVNC

Five variant classes were found to be significantly enriched solely in the LVNC cohorts, indicating such variants may yield a distinct non-compaction phenotype unrelated to either DCM or HCM. These include truncating variants in three cardiomyopathy-associated genes (*MYH7, PRDM16* and *ACTN2*) and specific variant classes in two arrhythmia-associated genes (*RYR2* and *HCN4*).

### MYH7 truncating variants

Overall, truncating variants in *MYH7* occur in 2.1% of LVNC cases compared to 0.05% in gnomAD (p= 2.4E-18) and are observed in each of the six constituent LVNC cohorts (Table 3, Table S6). These include a single splice donor variant, c.732+1G>A, present in six LVNC cases (with three other variants at this splice junction), as well as 10 other nonsense, frameshift and splice acceptor variants. Significant enrichment of *MYH7* truncating variants is observed even when excluding the c.732 splice junction variants (p=2.4E-09), which, along with the variant distribution throughout the *MYH7* transcript (Figure 3A), suggests that such variants are generally pathogenic in LVNC with haploinsufficiency as likely mechanism of action.

**Table 3:** Details of MYH7 truncating variants detected in LVNC cases in the six cohorts used in this meta-analysis.

| CDS | Protein | Population/Cohort | Cases | Clinical and family details |
| --- | --- | --- | --- | --- |
| *c.732 splice region* |  |  |  |  |
| c.732+1G>A | - | Netherlands | 2 | Variant detected in father, 10 year old son and new-born with LVNC, absent in 2 unaffected sisters of father<sup>27</sup>. |
|  |  | Switzerland/Germany | 2 | Variant in 6 affected family members, absent in 8 unaffected, LOD=2.55 (Family LVNC-101), variant in 3 affected family members, absent in 1 unaffected (Family LVNC-108). |
|  |  | Italy | 1 | Variant detected in 6 family members with varying degrees of non-compaction and/or hypertrabeculation (Figure 3). None had the c.732C>T common variant. |
|  |  | USA / LMM | 1 | Male, 0 months, cystic hygroma, Ebstein anomaly, LVNC and family history of LVNC. Variant also detected in twin brother with LVNC. Both had the c.732C>T common variant *in trans*. Both parents are unaffected. |
| c.732+1delG | - | USA / LMM | 1 | Female, 0 months, clinical diagnosis of LVNC with severely dilated left atrium and small secundum ASD, paternal family history of "enlarged heart" (father not tested) maternal family history of "fainting episodes" though variant not detected in mother. |
| c.732+3G>T | - | Italy | 1 | Detected in the affected mother (c.732C>T not detected) of an affected 5 year old son. |
| c.732+3G>C | - | Switzerland/Germany | 1 | Variant in 2 affected family members, absent in 1 unaffected (Family LVNC-109). |
| *Other MYH7 truncating variants* |  |  |  |  |
| c.745C>T | p.Arg249X | Poland/USA | 1 | - |
| c.798T>A | p.Tyr266X | Netherlands | 1 | Detected in girl, 4 years, and affected father (grandfather died suddenly at 60)<sup>27</sup>. |
| c.1903A>T | p.Lys635X | Poland/USA | 1 | - |
| c.2085_2097dup | p.Glu700Glnfs*37 | Netherlands | 1 | - |
| c.3100-2A>C | - | Netherlands | 1 | - |
| c.4125T>A | p.Tyr1375X | Netherlands | 1 | - |
| c.4354-2A>C | - | USA / LMM | 1 | Male, 15 years, clinical diagnosis of LVNC with biventricular dilation and PVCs |
| c.4588C>T | p.Arg1530X | France | 1 | - |
|  |  | USA / LMM | 1 | Female, 15 years, clinical diagnosis of LVNC. |
| c.5110C>T | p.Gln1704X | USA / LMM | 1 | Male, 0 months, with Ebstein anomaly and LVNC, family history of CHD (septal defect) and arrhythmia. Variant *in trans* with MYH7:p.Arg1897His. Both parents unaffected. |

The c.732+1G>A splice donor variant was detected in 6 individuals including two probands in the Dutch cohort^12^ (one of which was of Turkish origin^27^), two families in the Swiss/German cohort where it was determined by haplotype analysis to arise independently (reported by Klaassen *et al*. as c.818+1G>A^9^), one patient in the Italian cohort and one patient of unspecified ethnicity sequenced by LMM. These findings indicate that this is not a founder variant but one that has occurred recurrently in several different families and populations. This variant is detected in 1/125,745 individuals in the gnomAD exomes database (v2.1) and is therefore significantly enriched in the joint LVNC cohorts (Fisher’s exact test, p=5.1E-13). It is undetected in the gnomAD genomes (v3) database (71,702 samples).

In order to investigate this variant further we examined the Italian family pedigree in more detail. The proband is a 31-year old male with marked hypertrabeculation, apical and lateral non-compaction of the left ventricle and was diagnosed with LVNC at 14 years of age. He reported history of LVNC in his mother’s family and had undergone cardiac ablation at 15 years of age to treat ventricular pre-excitation. Targeted (re)sequencing using the Illumina TruSight Cardio panel alongside echocardiography and ECG was performed on the proband, his brother and 4 maternal family members. The variant was detected in all six family members, all showed varying degrees of myocardial hypertrabeculation (Figure 3D). We attempted to assess the effect of this variant on *MYH7* transcription using RNA from blood lymphocytes (other tissue was not available) but were unable to amplify any product (Supplemental Methods).

A deletion variant at the same splice donor base (c.732+1delG) and two other variants in the surrounding splice region (c.732+3G>T and c.732+3G>C [previously reported as c.818+3G>C^9^], outside the canonical splice site and not included in the burden analysis) were observed in three LVNC cases from different cohorts (Table 3, Figure 3B). All of these variants are predicted to affect splicing by the MaxEntScan algorithm^28^ (Figure 3C). No other variants in this splice region (intronic +1 to +8 bases) are detected in the gnomAD exomes (v2.1) or genomes (v3) databases. Interestingly, the adjacent synonymous exonic splice region variant, c.732C>T, is the only common splice region variant in *MYH7* (MAF=0.18) and is predicted by MaxEntScan to further disrupt splicing if co-occurring with the rare splice variants (Figure 3C). Whether there is a connection between the recurrence of rare splice variants in this splice region in LVNC cases and the presence of a common variant at the exon-intron boundary will require further investigation. However, no enrichment of the common splice region variant (c.732C>T) was observed in LVNC cohorts compared to population matched gnomAD controls (Table S7).

We then assessed NC/C ratios in population controls carrying *MYH7* truncating variants. Of 12 individuals in the UK Biobank and healthy volunteer cohorts with *MYH7* truncating variants, 6 had ratios greater than 2.3 (the diagnostic criteria for LVNC). The NC/C ratio was significantly greater in variant carriers compared to matched controls in these cohorts (2.7±1.2 vs 1.6±0.3, p=0.0034) (Figure 3E, Table S8).

### PRDM16 and ACTN2 truncating variants

The truncating variants in *PRDM16* (occurring in 1.4% of LVNC cases) include two variants previously published in the Swiss/German cohort^22^, three variants in the Dutch cohort^12^ and one variant in the LMM cohort (Table S9). Three truncating variants in *ACTN2* were observed in the Dutch cohort and one variant, p.Arg192X, in the Italian cohort. These findings add to existing evidence (Table 4 and Discussion) for a role for *PRDM16* and *ACTN2* in LVNC. Notably, there is evidence for constraint of truncating variants in gnomAD for both of these genes with fewer observed than expected variants for *PRDM16* (o/e ratio = 0.08) and *ACTN2* (0.12) and both defined as loss-of-function intolerant (pLI=1), offering additional supportive evidence for the deleteriousness of these variant classes.

**Table 4:**
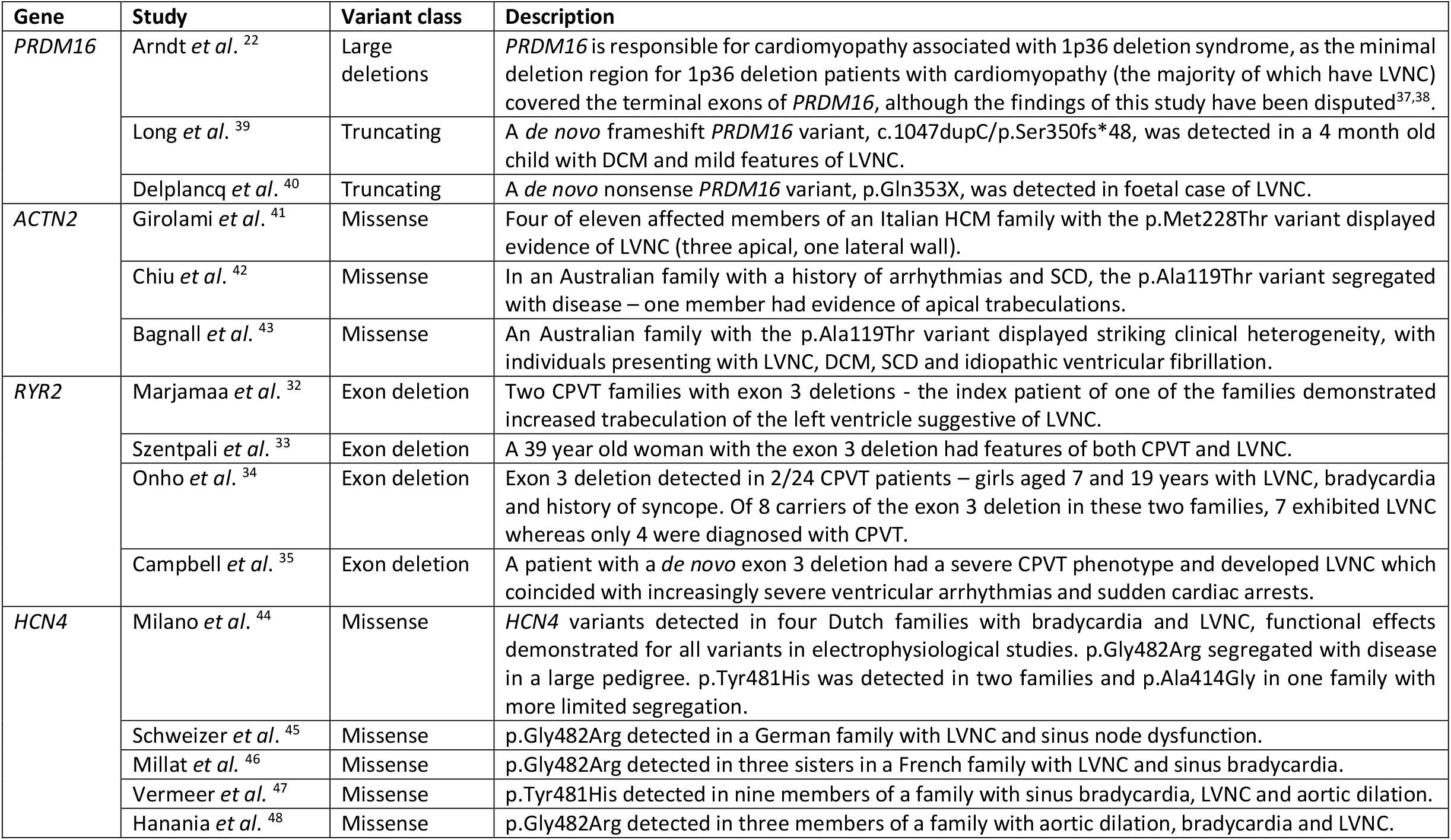
Summary of published studies supporting a role in the non-compaction phenotype for PRDM16 and ACTN2 and in arrhythmogenic and non-compaction phenotypes for RYR2 and HCN4.

### Variants in arrhythmia-associated genes - RYR2 and HCN4

Variants in *RYR2* are the primary cause of catecholaminergic polymorphic ventricular tachycardia (CPVT), with pathogenic missense variants present in approximately 50% of cases. Deletion of exon 3 in *RYR2* has been described in CPVT cases^30,31^ and recently in a number of patients and family pedigrees with complex phenotypes that include LVNC and CPVT^32–35^ (Table 4). In this meta-analysis, five SVs, corresponding to deletions of whole exons, were observed in the *RYR2* gene in LVNC cases (three of exon 3 and one each of exons 2 and 19) (Table S9). No exon deletions for *RYR2* were detected in the gnomAD SV database derived from whole genome sequencing of 14,891 individuals^36^, suggesting such variants are rarely observed in the population.

The overall enrichment of non-truncating variants in *HCN4* (3.7% vs 1.1%) did not meet the significance threshold with multiple testing correction (p=0.002). However, we found significant clustering of non-truncating variants in the trans-membrane region of the ion channel encoded by *HCN4* (p=3.5×10^−7^, Table S3) and therefore performed burden testing for variants within and outside this region. While no enrichment occurred outside of the transmembrane region (p=0.87), a significant excess was observed for variants within the transmembrane region (3.3% vs 0.1%, p=4.8E-09). Of the *HCN4* carriers described in the constituent studies of this meta-analysis, a patient with the p.Tyr481His variant initially presented with bradycardia (although this patient also had two other variants classified as pathogenic - *FKTN*:p.Tyr371Cys and *RBM20*:p.Tyr283Glnfs*14)^12^ while 3/5 proband carriers of *HCN4* variants in the French study were reported to have bradycardia, as did four carriers of the p.Gly480Cys variant in one family^13^ (Table S9). In contrast, the Italian LVNC patient with the only non-transmembrane *HCN4* variant detected in this study (p.Gly1077Ser) had a normal ECG, suggesting this variant is unlikely to be disease-causing.

## Discussion

The meta-analysis of genetic sequencing data from 840 cases described here provides much needed clarity concerning the genetic basis of LVNC. By amalgamating data from several recently published and moderately sized studies with two new cohorts, we were able to identify genes and variant classes with robust statistical evidence of association with LVNC. These findings highlight the diverse aetiology underlying this phenotype and inform how genetic testing should be applied and interpreted for patients presenting with LVNC.

Our results also reveal a substantial overlap in genes and variant classes enriched in LVNC with those of the more genetically well-defined cardiomyopathies of DCM and HCM. Several other genes associated with DCM/HCM were nominally enriched in LVNC cases but did not meet the significance threshold (e.g. truncating variants in the *DSP* DCM gene and non-truncating variants in the *MYL2* HCM gene, Table S5). This likely reflects the more heterogeneous aetiology of LVNC, suggesting that genes more rarely implicated in DCM/HCM are generally causative in an even smaller proportion of LVNC cases.

These findings are consistent with the increasingly held view that LVNC largely belongs to the spectrum of more common and established cardiomyopathies. The expression of the particular trait (i.e. hypertrabeculation) represents a striking phenotypic variation but ultimately one with limited impact on the pathophysiology and natural history of the underlying paradigm (HCM or DCM). Such a concept is well reflected in recent management consensus documents which recommend assessing risk and treating LVNC according to the principles of HCM or DCM, as appropriate. The factors that cause patients with DCM/HCM-causing variants to develop and/or present with LVNC rather than the more standard phenotypes are unknown but could involve genetic and non-genetic modifiers. One intriguing candidate is the *MIB1* gene, a regulator of the Notch signalling pathway that has previously been implicated in LVNC^49^. Three *MIB1* truncating variants were identified in the Dutch LVNC cohort, and in all cases occurring together with a *TTN* truncating variant, suggesting they could modify the DCM phenotype typically associated with *TTN* variants^12^. As *MIB1* truncating variants are relatively common in the population, with the tenth lowest pLI score of all human genes in gnomAD and an o/e ratio of 1.83 (1.54-1.97), they may act as modifiers rather than primary pathogenic variants.

A recent study assessing clinical and genetic screening in families of LVNC patients provided further evidence for this hypothesis — many family members had DCM or HCM without non-compaction, and the genotype of the proband was broadly predictive of the phenotype in relatives (*TTN* and *MYH7* tail domain variants were associated with DCM in relatives and *MYBPC3* variants were associated with HCM)^50^. Our results suggest it would be prudent to include all proven DCM and HCM genes for genetic testing in LVNC cases (whether these genes have been validated though statistical association^18,20^ or the curation of published evidence^17,20^) in order to inform clinical management in both patients and family members through clinical and genetic cascade screening.

In contrast, van Waning *et al*. also found that *MYH7* head domain variants in probands were predictive of isolated LVNC in relatives, indicating that specific variant classes may be associated with a distinctive non-compaction phenotype rather than underlying DCM/HCM. Accordingly, we also detected additional variant classes not associated with other cardiomyopathies and enriched in LVNC patients in 5-10% of cases. This patient subset may therefore have an aetiology separate from other cardiomyopathies or cardiac conditions and represent genetically distinct disease where non-compaction is the primary or presenting phenotype.

Of the variant classes unique to LVNC, perhaps the most notable are truncating variants in *MYH7*. Such variants have generally been considered as non-pathogenic and indeed are not associated with either HCM or DCM^18^, where non-truncating (largely missense) variants act through a dominant negative mechanism (with opposing activating and inactivating functions). However, they are observed in over 2% of LVNC cases, consistently across all of the constituent cohorts in this meta-analysis, and are significantly enriched over the general population rate in gnomAD (p=2.4E-18). Data from population controls provide further evidence for the role of *MYH7* truncating variants in LVNC. A high population-level penetrance was observed for these variants, with 50% of carriers meeting the diagnostic criteria for LVNC. NC/C ratios were also significantly greater in carriers compared to matched controls. The exact mechanism of action of these variants remains to be fully determined although the distribution of truncating variants throughout the *MYH7* gene would support nonsense mediated decay and haploinsufficiency. The large number of variants clustering around one splice region is particularly intriguing given that the most common of these, c.732+1G>A, does not appear to be a founder mutation. More research will be required to establish why variants in this specific location are particularly associated with LVNC.

Truncating variants in two other cardiomyopathy-associated genes, *ACTN2* and *PRDM16*, also appear to be associated solely or largely with an LVNC phenotype albeit in a small proportion of cases (approximately 1% each). These observations are supported by other associations with LVNC at these loci (Table 4), e.g. the LVNC phenotype underlying 1p36 deletion syndrome that may involve *PRDM16*^22^. A role for *PRDM16* in DCM has also been postulated. However, missense variants detected in a small DCM cohort were at a frequency (2.3%) equivalent to the gnomAD population rate^22^, although a putative *de novo* truncating variant was recently described in a paediatric patient with DCM and mild features of LVNC^39^. Although no excess of *ACTN2* rare variants (truncating or non-truncating) have been observed in DCM or HCM cohorts^18,20^, two missense variants have been reported in pedigrees with complex heterogeneous phenotypes that include non-compaction (Table 4)^41–43^. The significant association of *ACTN2* truncating variants with LVNC described here may indicate that such loss of function variants in this gene lead to more overt presentation of LVNC.

This study has shown that prior reports of *RYR2* and *HCN4* variants with LVNC and CPVT or bradycardia, respectively, are supported by statistically significant associations in case-control cohort analysis. Based on the limited data available thus far, their role in LVNC seems to be limited to very specific variant classes in these genes. The pathogenicity of the *RYR2* exon 3 deletion has now been established in several reports (Table 4), but the deletions of two other *RYR2* exons (2 and 19) described here suggests a potentially broader role for this variant class in LVNC (although the pathogenicity of these novel variants is yet to be unambiguously established). The enrichment of missense variants in the *HCN4* transmembrane region is also consistent with previous reports describing combined non-compaction and arrhythmia phenotypes (Table 4). Although the published reports on these variant classes reveal considerable phenotypic heterogeneity, it is conceivable that such patients could present primarily with an LVNC phenotype. Detecting such variants with LVNC genetic testing panels could identify patients (or family members carrying the same variants) at risk of arrhythmogenic events (particularly the severe events associated with CPVT).

While association has been demonstrated here for both established cardiomyopathy genes and variant classes specifically associated with LVNC, no significant excess of rare variation was observed for the majority of genes sequenced in the constituent studies, similar to previous findings for HCM^20^ and DCM^18^. While a lack of excess does not necessarily preclude a role in disease, it does indicate that variants in such genes are likely to be, at best, very rarely causative. Future studies with larger LVNC cohorts may be sufficiently powered to detect a significant case excess for genes more rarely involved in the disease, especially for variant classes with proven association to other cardiomyopathies as described above. However, these results again highlight the dangers of sequencing, in the diagnostic context, broad panels of genes that include many not fully validated for the disease in question, increasing the uncertainty associated with results (more variants of unknown significance) as well as the risk of false positive results. As with other conditions, clinical genetic testing for LVNC cases should be restricted to those genes with a proven association to disease.

### Limitations

There are some limitations associated with the analysis described in this study. As this is a meta-analysis of 6 different LVNC cohorts, there may be minor differences in how LVNC was diagnosed between the different studies and in the inclusion or exclusion criteria for patients with other cardiac phenotypes in addition to LVNC. However, we do observe broad consistency across cohorts for the significantly associated variant classes (e.g. *MYH7* truncating variants, Table S6), despite the limited cohort sizes, indicating this is not a major confounding factor. The burden analysis here also compares sequencing data from different studies and platforms which may introduce bias when comparing cases to reference population samples. As described previously, we adjusted for expected poorer coverage in the WES data of gnomAD^18^ and used filtering allele frequency values in gnomAD to define rarity, so to minimize any confounding effects due to population stratification. Our previous work for HCM showing strong correlation between genes validated through this rare variant burden approach^18,20^ and those validated by the curation of published evidence^17,20^ demonstrates the robustness of this approach for identifying the most relevant causative genes in Mendelian diseases. Future larger single-centre studies, or coordinated efforts between different centres that synchronise diagnostic criteria and sequencing methods will be valuable in confirming the gene associations described here and clarifying any role for other genes and variant classes in LVNC.

### Conclusion

We have demonstrated a significant association of a number of gene and variant classes with LVNC in this meta-analysis of cohort sequencing data. These findings confirm a large genetic overlap between LVNC and DCM, HCM and arrhythmogenic diseases and provide strong evidence to support the hypothesis that many LVNC cases are a variable morphological phenotype of an underlying cardiac disease. However, we have also identified a distinct genetic aetiology in a subset of LVNC cases, with variant classes such as *MYH7* truncating variants that are not associated with other cardiac conditions, and confirmed the association of *RYR2* and *HCN4* with complex non-compaction/arrhythmia phenotypes. This data suggests that LVNC is a heterogeneous condition that can be associated with cardiomyopathies, arrhythmias, other underlying cardiac and syndromic disease as well as potentially acting as an isolated and genetically distinct pathology. Our results indicate that focused genetic testing in patients that present with LVNC may distinguish between these different aetiologies, inform clinical management for patients and their relatives and help to distinguish pathological from physiological non-compaction. This study also demonstrates the power of statistically robust genetic association studies in characterising and classifying complex clinical phenotypes.

## Data Availability

All data is available in the supplemental material.

